# Early Pregnancy Glycosylated Hemoglobin in Relation to Gestational Diabetes Mellitus and Adverse Pregnancy Outcomes: Evidence from a Multisite Low- and Middle-Income Countries Cohort

**DOI:** 10.64898/2026.09.23.26362980

**Authors:** Wen-Chien Yang, Sarah Sunil Joseph, B Vijayalekshmi, Bethany L Freeman, Muhammad Imran Nisar, Qing Pan, Santosh Joseph Benjamin, Jayakumar Amirtharaj G, Neeraj Sharma, Sarmila Mazumder, Zahra Hoodbhoy, Victor Akelo, Florence Aweyo, Emily R Smith, Christopher N Mores, Erin M Oakley, M Bridget Spelke, Humphrey Mwape, Sarah Mukuka, Margaret P Kasaro, Anne George Cherian, the Pregnancy Risk, Infant Surveillance, and Measurement Alliance (PRISMA) Consortium

## Abstract

**Background:** Oral glucose tolerance test (OGTT), the gold standard for diagnosing gestational diabetes mellitus (GDM), faces significant implementation challenges, prompting interest in alternative biomarkers, such as glycated hemoglobin (HbA1c), for early detection. This study aimed to assess the association between early pregnancy HbA1c and subsequent GDM and adverse perinatal outcomes in low- and middle-income countries.

**Methods:** We analyzed data from the **Pregnancy Risk, Infant Surveillance, and Measurement Alliance - Maternal and Newborn Health** (PRISMA-MNH) cohort, which followed pregnant women enrolled before 20 weeks of gestation across five study sites in Kenya, India, Pakistan, and Zambia. We excluded participants with overt diabetes, severe anemia (Hb <7.0 g/dl), multiple gestations, or pregnancies that ended before 24 weeks of gestation. HbA1c was measured at enrollment. The primary outcome was GDM, diagnosed according to the International Association of Diabetes and Pregnancy Study Groups (IADPSG) criteria based on OGTT performed after 24 weeks of gestation. Secondary outcomes included hypertensive disorder of pregnancy, emergent Cesarean section, preterm birth, large for gestational age, and stillbirth after 28 weeks. We used AUC and ROC analyses to evaluate predictive performance and single- and two-threshold approaches to identify optimal cutoffs for predicting GDM. We used modified Poisson regression with robust standard errors to assess associations between early HbA1c and outcomes, and meta-analysis to pool site-specific estimates.

**Results:** Among 10,918 pregnancies, GDM prevalence was 6.4%. Women with GDM were older (28.0±5.4 versus 26.0±5.3 years) and had higher BMI at enrollment (25.0±5.8 versus 22.9±5.0 kg/m^2^) than women without GDM. The AUC for HbA1c alone in predicting GDM was 0.65 (95% confidence interval 0.63–0.67). A single early HbA1c threshold of 5.4%, based on the largest Youden’s index, showed limited predictive performance (sensitivity 46.7%, specificity 75.8%). A two-threshold approach identified rule-out and rule-in cutoffs of 5.0% (sensitivity 80.3%, specificity 35.6%) and 5.5% (sensitivity 34.5%, specificity 83.1%), respectively. In adjusted models, each 0.1% increase in HbA1c was associated with a 12% higher risk (RR 1.12, 95% CI 1.10–1.15) of GDM, while no significant associations were observed for any secondary outcomes.

**Conclusion:** Early pregnancy HbA1c is independently associated with an increased risk of subsequent GDM. However, it demonstrated limited discriminatory ability as a standalone alternative to OGTT for identifying women who would later develop GDM. The identified rule-out threshold may help identify women at low risk of GDM, although the majority would still require OGTT for definitive diagnosis.

## Introduction

Gestational diabetes mellitus (GDM), defined as glucose intolerance first recognized during pregnancy [1], is an increasingly prevalent global health concern. Current estimates suggest that GDM affects approximately 14% of pregnancies globally, with reported prevalence ranging from 5% to 27.6% depending on the population and diagnostic criteria used.[2, 3] The burden is disproportionately concentrated in low- and middle-income countries (LMICs), which account for more than 90% of cases.[4] The IDF Diabetes Atlas reported standardized GDM prevalences of 20.8% in South-East Asia and 27.6% in the Middle East and North Africa. [3] A substantial body of evidence links GDM with adverse pregnancy and birth outcomes, as well as an increased lifetime risk of type 2 diabetes and cardiovascular diseases in both mothers and their offspring [5, 6], highlighting the importance of early detection for timely intervention and risk reduction.

International guidelines from the World Health Organization (WHO) and the International Association of Diabetes and Pregnancy Study Groups (IADPSG) recommend the oral glucose tolerance test (OGTT) for diagnosing GDM at 24 to 28 weeks of gestation.[7] Although OGTT remains the gold standard [8], its implementation in low-resource settings is often constrained by limited laboratory capacity, shortages of trained personnel, inadequate access to health facilities, and broader challenges in delivering routine antenatal care.[9, 10] For pregnant women, the test is logistically demanding, requiring overnight fasting, ingestion of a glucose load, multiple blood draws, and prolonged clinic visits. These requirements can be particularly burdensome for women traveling long distances or managing competing responsibilities.[11, 12] Given these constraints, the WHO has emphasized the need for simplified, context-appropriate, and point-of-care approaches to facilitate GDM identification in resource-limited settings.[13]

Glycated hemoglobin (HbA1c) has been proposed as a potential biomarker for early detection of GDM [4], as it reflects average glycemia over the preceding 2-3 months and does not require fasting and multiple blood draws. However, significant evidence gaps remain. First, meta-analyses suggest that early pregnancy HbA1c, while promising, has high specificity (75–85%) but low-to-modest sensitivity (40–60%) for predicting GDM, with substantial heterogeneity across populations. This means that HbA1c, when used as a standalone diagnostic tool, misses a large proportion of true GDM cases.[14, 15] Second, there is limited evidence on using early pregnancy HbA1c to predict GDM in diverse low-resource settings where anemia, nutritional deficiencies, hemoglobinopathies, and regional differences may affect HbA1c interpretation. The STRiDE study, a large multisite cohort of pregnant women in India, Kenya, and the UK, found that early HbA1c alone or as part of a composite risk score with age, BMI, and family history of diabetes could reduce the need for OGTT by 50 to 64%.[4] However, aside from this study, most existing evidence is derived from ethnically homogeneous populations and uses non-harmonized diagnostic criteria for GDM, limiting broader public health implications across diverse settings.[14–17]

To address these evidence gaps, this study aimed to assess the predictive performance of early pregnancy HbA1c for identifying GDM according to the IADPSG criteria, and to examine the associations between early pregnancy HbA1c and major maternal and perinatal outcomes.

## Methods

### Setting and study population

We analyzed data collected from September 2022 through January 2026 as part of the Pregnancy Risk, Infant Surveillance, and Measurement Alliance (PRISMA) Maternal and Newborn Health (MNH) study. The PRISMA MNH study is a cohort study in six sites of five low-resource countries (Kintampo, Ghana; Kisumu, Kenya; Lusaka, Zambia; Karachi, Pakistan; Vellore, southern India; and Hodal, northern India), designed to identify risk factors of maternal and infant morbidity and mortality by collecting longitudinal data from early pregnancy to the postpartum period [18, 19] (ClinicalTrials.gov: NCT05904145). All sites followed a harmonized study protocol and standard operating procedures for data collection, where participants were enrolled consecutively via community- and facility-based surveillance, at slightly different times across sites (September 2022, Pakistan; November 2022, Kenya; December 2022, Zambia and Ghana; July 2023, South India; December 2023, North India). Pregnant women were enrolled before gestational age (GA) 20 weeks and assessed at GA 20, 28, 32, and 36 weeks, at delivery, and up to 1 year postpartum.

This analysis excluded participants with overt diabetes (defined as participant-reported or medical records indicating pre-existing diabetes or an HbA1c ≥6.5% at enrollment) and those with severe anemia at enrollment (defined as hemoglobin <7 g/dL), because anemia might falsely elevate or lower HbA1c levels depending on the etiology. Twin- or triplet-pregnancies were excluded. While our prespecified acceptable GA window for OGTT was 26 to 30 weeks, we included OGTTs done after GA 24 weeks in accordance with international guidelines recommending an OGTT between GA 24 and 28 weeks. Thus, pregnancies that ended before 24 weeks were excluded because the primary outcome, GDM, would not be assessed until GA 24 weeks. This analysis excluded data from the PRISMA Ghana site because we identified challenges with consistent HbA1c measurement through participation in an ongoing external quality assurance program. As a result, the quality of HbA1c data from this site could not be adequately verified.

### Data collection and outcomes of interest

In the PRISMA MNH study, HbA1c was measured early in pregnancy at study enrollment (before GA 20 weeks) across sites using a harmonized protocol (HPLC, Bio-rad D-100 for South India; HPLC, Tosoh for North India; ECLIA, Roche Cobas Pure for Pakistan; HPLC, Biorad D10 Hemoglobin testing system for Kenya; turbidimetric inhibition immunoassay and Roche Cobas C111 for Zambia). The primary outcome of this analysis is GDM, diagnosed by an OGTT based on the IADPSG criteria, requiring three blood glucose measurements: fasting and one and two hours after oral ingestion of a 75g glucose solution. GDM was diagnosed if any of the following criteria were met: a fasting glucose level ≥ 5.1 mmol/L, a 1-hour OGTT glucose level ≥ 10.0 mmol/L, or a 2-hour glucose level ≥ 8.5 mmol/L.[7] The North India site did not measure 1-hour OGTT glucose. In this analysis, participants were classified as not having GDM if all three blood glucose measurements (or two measurements at the North India site) were below the criteria; conversely, participants were classified as having GDM if at least one measurement met or exceeded the criteria, regardless of whether the remaining measurements were below the criteria or missing. Blood glucose levels were measured using harmonized methods across sites (Chemistry analyzers (colorimetric methods), Beckman AU5800 for South India; Chemistry analyzers (colorimetric methods), Roche Cobas Pro for North India; Electrochemiluminescence (photometric end point), Roche Cobas pure for Pakistan; Zybio Chemistry Analyzer, Model EXC 200 for Kenya; Chemistry analyzer (photometric), Roche Cobas C111 for Zambia. All sites followed a harmonized study laboratory manual specifying sample handling, and all site laboratories were enrolled in an external Quality Assurance Scheme (EQAS) through the College of American Pathologists (CAP) and underwent proficiency testing three times per year, with results reviewed centrally by the PRISMA MNH lab coordinating team to verify accuracy and inter-laboratory comparability. Because HbA1c testing (<20 weeks GA) and OGTT (≥24 weeks GA) were performed sequentially, personnel administering and interpreting the OGTT were naturally blinded to the early HbA1c research values, and diagnostic classifications were evaluated independently. Any indeterminate HbA1c results or unreadable OGTT samples were treated as missing data and excluded from the primary outcome classification.

The secondary outcomes included hypertensive disorder of pregnancy (HDP, defined as having hypertension at or after 20 weeks of gestation, including gestational hypertension, preeclampsia with and without severe features), emergent Cesarean section, preterm birth (defined as birth before GA 37 weeks), large for gestational age (LGA, defined as newborn with a birth weight > 90th percentile according to the INTERGROWTH-21st standard) [20], and stillbirth after GA 28 weeks.

### Statistical analysis

The sample size was determined a priori by the parent PRISMA MNH cohort’s enrollment targets, which were powered to detect risk factor associations as described in the study protocol.[18] We first summarized participant characteristics by GDM status, including age and age groups (<20 years old, 20 to 34 years old, and ≥35 years old), anthropometric measurements (weight, height, and body mass index (BMI)) and BMI categories (underweight: BMI <18.5; normal: BMI 18.5 to <25: overweight: BMI 25 to <30; obese: BMI ≥30) at enrollment, and education level. We also described pregnancy characteristics and medical history, including multiparity, smoking history, prior GDM, and prior macrosomia, if any. The distribution of early HbA1c was examined by study site and GDM status.

Second, we evaluated the predictive performance of early HbA1c for identifying GDM using receiver operating characteristic (ROC) curves and area under the curve (AUC) analyses in both pooled and site-specific datasets. The Delong method was used to estimate the 95% confidence interval (CI) of AUC. To determine optimal early HbA1c cutoffs, we adopted both single- and two-threshold approaches. The single-threshold approach identified the cutoff that best distinguished GDM from non-GDM cases using either the largest Youden’s index or the smallest Euclidean distance. The two-threshold approach identified a lower cutoff with 80% sensitivity to rule out GDM and a higher cutoff with 80% specificity to rule in GDM. We reported sensitivity, specificity, positive and negative predictive values (PPV, NPV), true positives (TP), false positives (FP), true negatives (TN), false negatives (FN), and accuracy where appropriate.

Third, we estimated the associations between early HbA1c, as a continuous predictor, and each outcome using three modified Poisson regressions with robust standard errors for each study site separately. The assumptions for modified Poisson regressions with robust standard errors were assessed in advance, including binary outcomes, adequate sample sizes, independence of observations, etc. Model 1 included only early HbA1c; model 2 adjusted for maternal age groups, BMI categories, years of education, multiparity, and gestational age at HbA1c measurement; the covariates were selected from a literature review.[4, 21–24] Model 3 was restricted to those with prior pregnancies and additionally adjusted for a past history of GDM status. Model 2 was designated a priori as the primary model for association testing; model 3 was secondary and exploratory. Site-specific relative risks (RR) were pooled to generate an overall effect measure using a random-effects meta-analysis approach.[25] Several sensitivity analyses were conducted, including one stratified by anemia status (no anemia versus mild and moderate anemia), one among those with a normal early HbA1c value (< 5.7%), and one excluding those with HbA1c measured after GA 18 weeks. We also examined how the rule-out and rule-in cutoffs that were identified through the two-threshold approach, treated as dichotomous predictors, were associated with adverse outcomes, adjusting for covariates in model 2.

Lastly, we used generalized additive models (GAMs), adjusted for covariates in model 2, to explore potential nonlinear relationships between continuous early HbA1c and adverse outcomes. Throughout this paper, a complete-case analysis was performed, indicating that observations with missing data on covariates were dropped from the models. No missing data imputations were performed. All analyses were performed using R statistical software (version 4.4.3; R Foundation for Statistical Computing, Vienna, Austria; packages: sandwich, metafor, pROC, ect).

## Results

### Participant characteristics and HbA1c distribution

Among the 10,918 participants included in this analysis **(Figure 1)**, 696 (6.4%) were diagnosed with GDM based on the IADPSG criteria **(Table 1)**. The GDM group was older, with 12.8% aged 35 or older, versus 8.2% in the non-GDM group. Women with GDM had higher weight at enrollment than those without GDM (weight 60.3±15.4 kg versus 56.1±13.4 kg) and were more likely to be overweight or obese at enrollment (46.2% versus 28.7%). Among multiparous women, the GDM group had a higher proportion with a history of GDM versus the non-GDM group (4.2% versus 0.9%, respectively). HbA1c in early pregnancy was slightly higher among those with GDM (5.3 ±0.4% versus 5.1±0.4%). Across all sites, women diagnosed with GDM had higher early HbA1c levels than those without GDM **(Figure 2)**. The mean gestational age was 13.1 weeks (standard deviation (SD) 3.6) at HbA1c measurement and 27.5 weeks (SD 1.2) at OGTT. Details on the OGTT timing are provided in **Supplemental Figure 1** and **Supplemental Table 1**. Maternal characteristics and HbA1c statistics stratified by site and by GDM status were presented in **Supplemental Table 2**. Characteristics of participants with and without sufficient OGTT data to determine GDM status were also shown in **Supplemental Table 3**.

**Figure 1.**
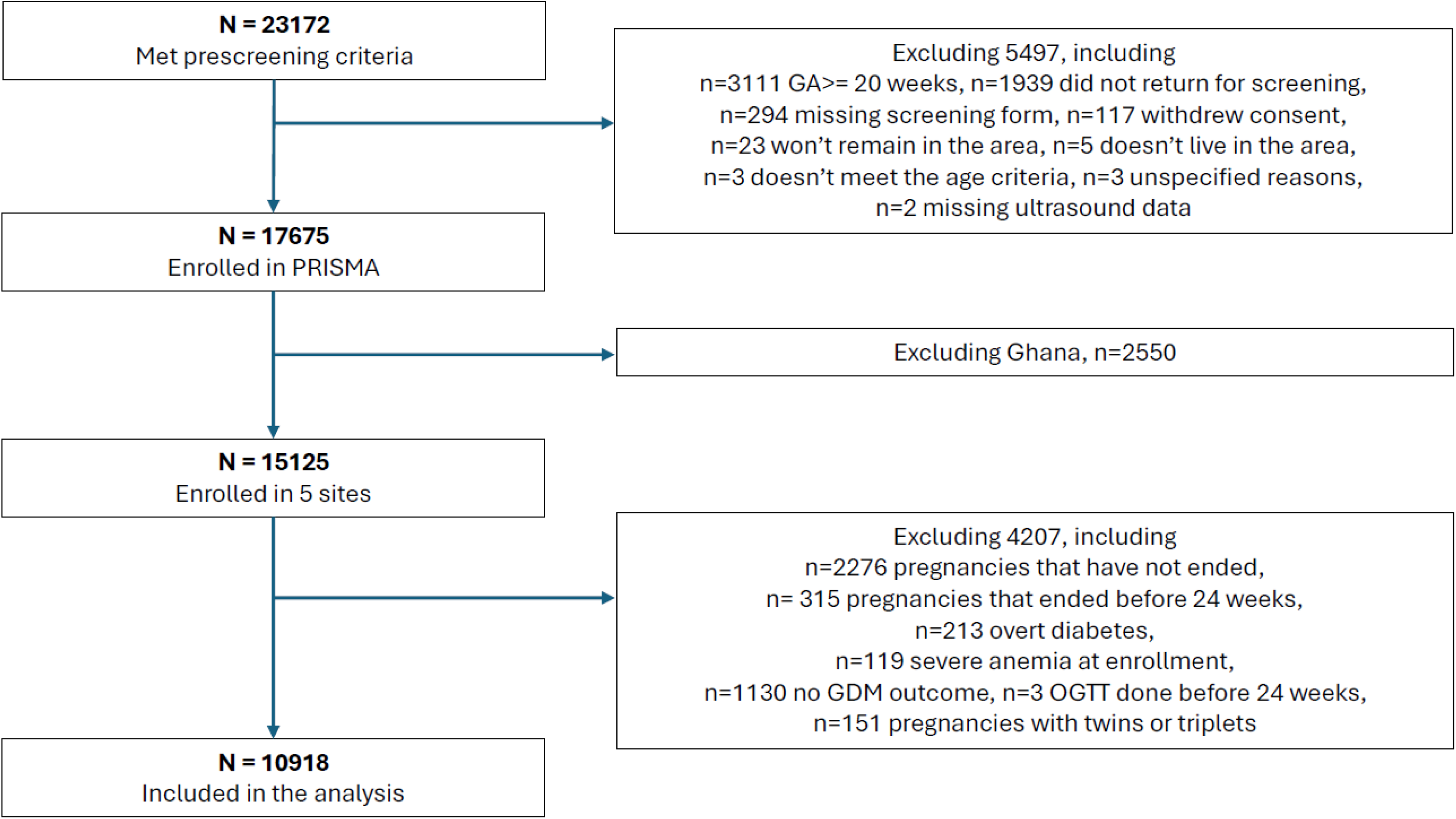
Flowchart of participant enrollment.

**Figure 2.**
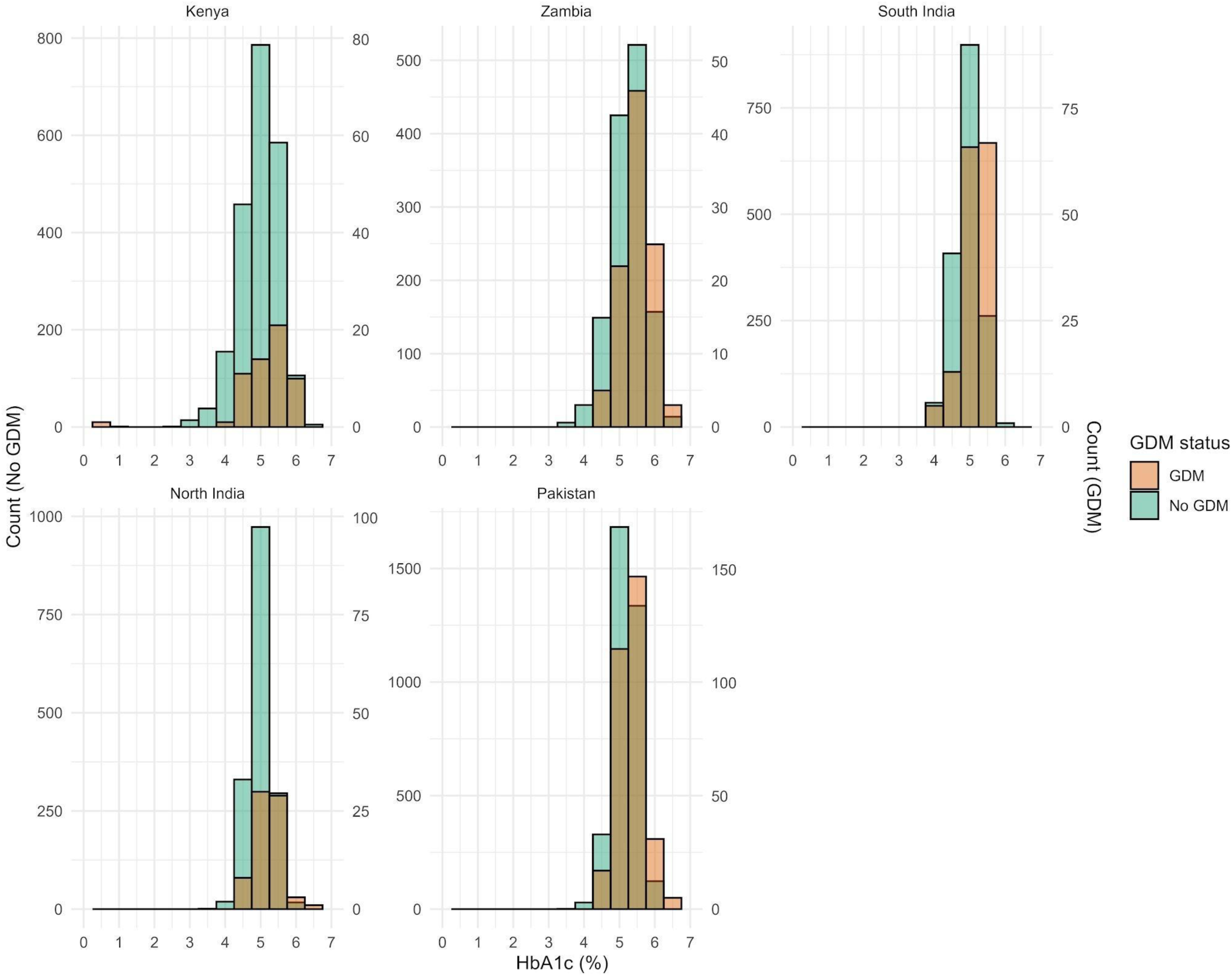
Early HbA1c distribution by site and GDM status.

**Table 1.**
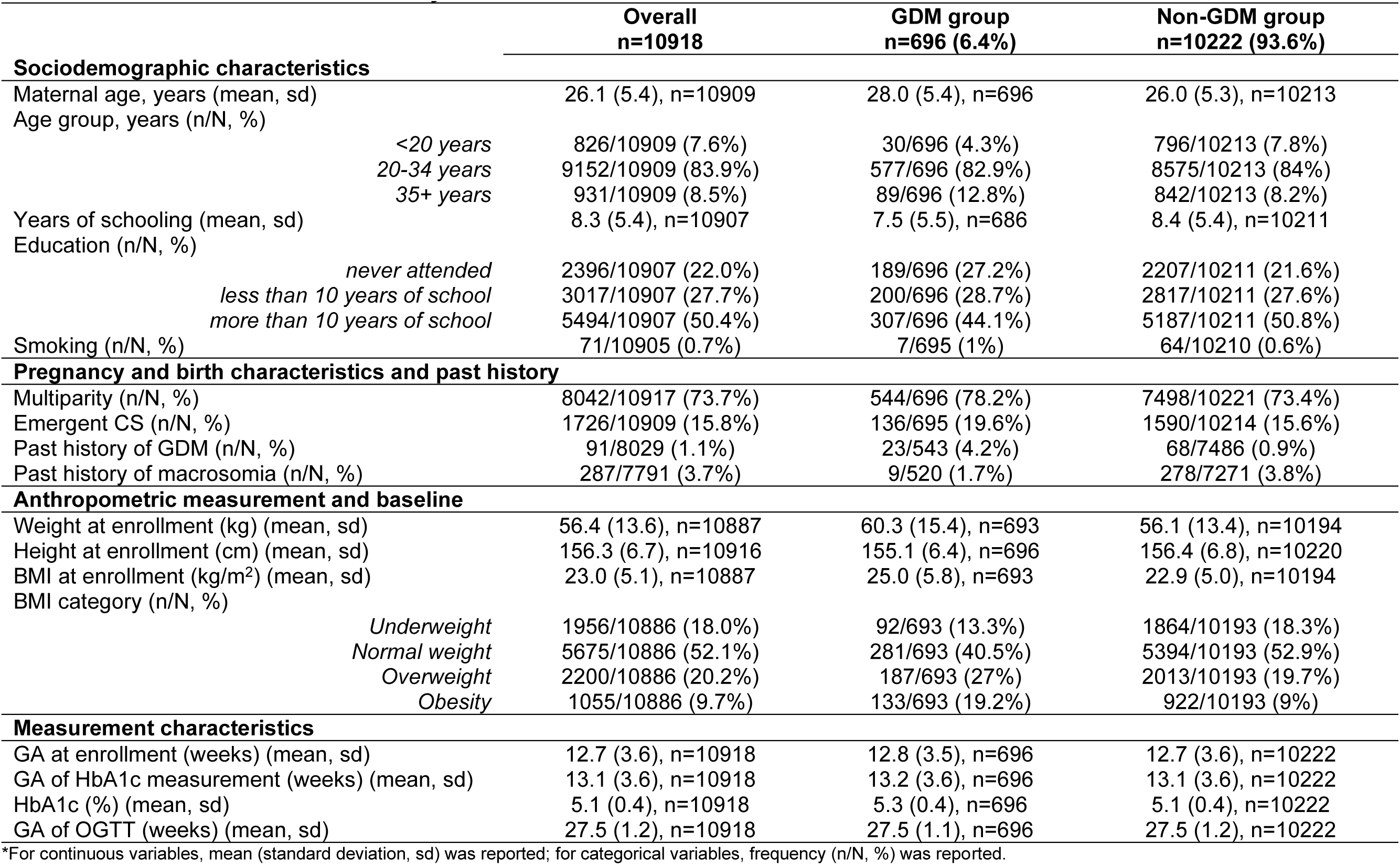
Maternal characteristics by GDM status.

### ROC and AUC analyses

Using the single-threshold approach, the optimal HbA1c cutoff was 5.4% (sensitivity 46.7%, specificity 75.8%) based on the largest Youden’s index and 5.3% (sensitivity 55.7%, specificity 66.5%) based on the smallest Euclidean distance **(Table 2)**. However, both cutoffs had low sensitivities (46.7% for HbA1c 5.4%; 55.7% for HbA1c 5.3%) and high false positive rates (24.2% for HbA1c 5.4%; 33.5% for HbA1c 5.3%). Across sites, HbA1c cutoffs varied, ranging from 5.1% in South India, based on the smallest Euclidean distance, to 5.5% in Zambia, based on both the largest Youden’s index and the smallest Euclidean distance **(Supplemental Table 4)**. Early HbA1c alone had a moderate ability to distinguish participants who would later develop GDM from those who would not, with an AUC of 0.65 (95% CI 0.63–0.67) **(Figure 3)**. The AUC varied slightly by site, ranging from 0.61 (95% CI 0.53–0.69) in Kenya to 0.69 (95% CI 0.64–0.73) in South India.

**Figure 3.**
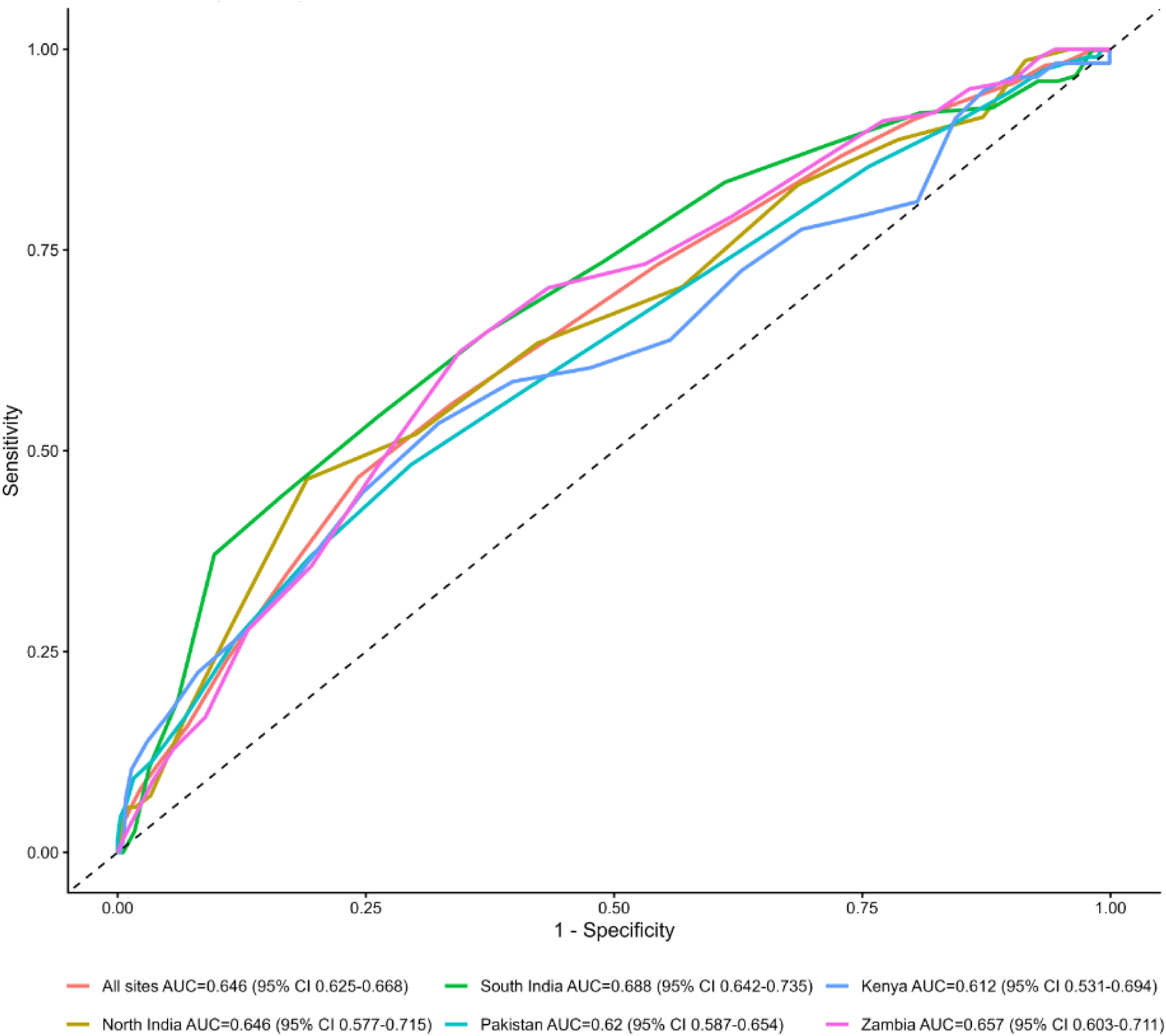
Receiver operating characteristic (ROC) curves of early HbA1C in predicting GDM, stratified by study site.

**Table 2.**
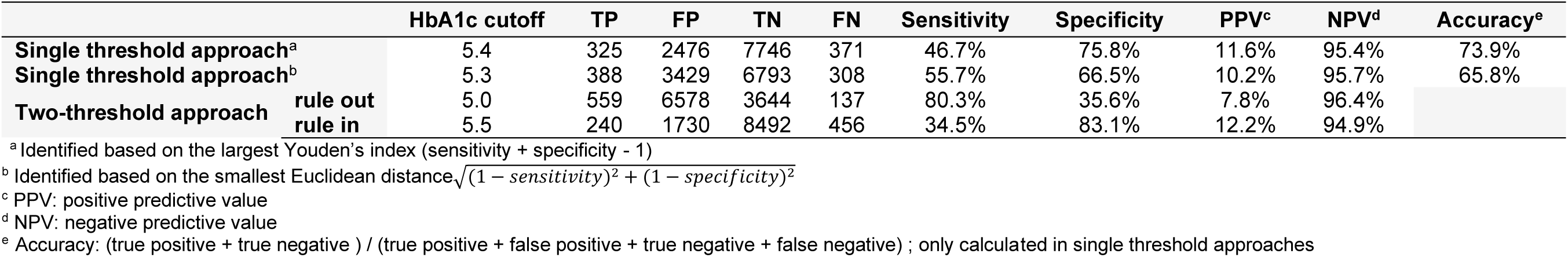
Early HbA1c cutoffs for identifying GDM using a single-threshold and two-threshold approaches.

In the two-threshold approach, a HbA1c value of 5.0% was the lower cutoff for ruling out GDM (sensitivity 80.3%, specificity 35.6%), while a value of 5.5% was the higher cutoff for ruling in GDM (sensitivity 34.5%, specificity 83.1%) **(Table 2)**. Across sites, Zambia had the highest rule-out and rule-in cutoffs at 5.2% and 5.7%, respectively **(Supplemental Table 4)**.

### Associations between early HbA1c, GDM, and adverse outcomes

Each 0.1% increase in early HbA1c was associated with a 12% higher risk of GDM (RR 1.12, 95% CI 1.10–1.15; p<0.001) in the adjusted model, and the association was similar in the analysis restricted to multiparous women (RR 1.12, 95% CI 1.10– 1.15; p<0.001) **(Figure 4** and **Supplemental Table 5)**. No significant associations were observed for the secondary outcomes in model 2, although a borderline significance was noted for the outcome of emergent Cesarean section. In model 3 restricting to multiparous women, a significant association with emergent Cesarean section, with a borderline association for HDP. Site-specific estimates were provided in **Supplemental Table 6**.

**Figure 4.**
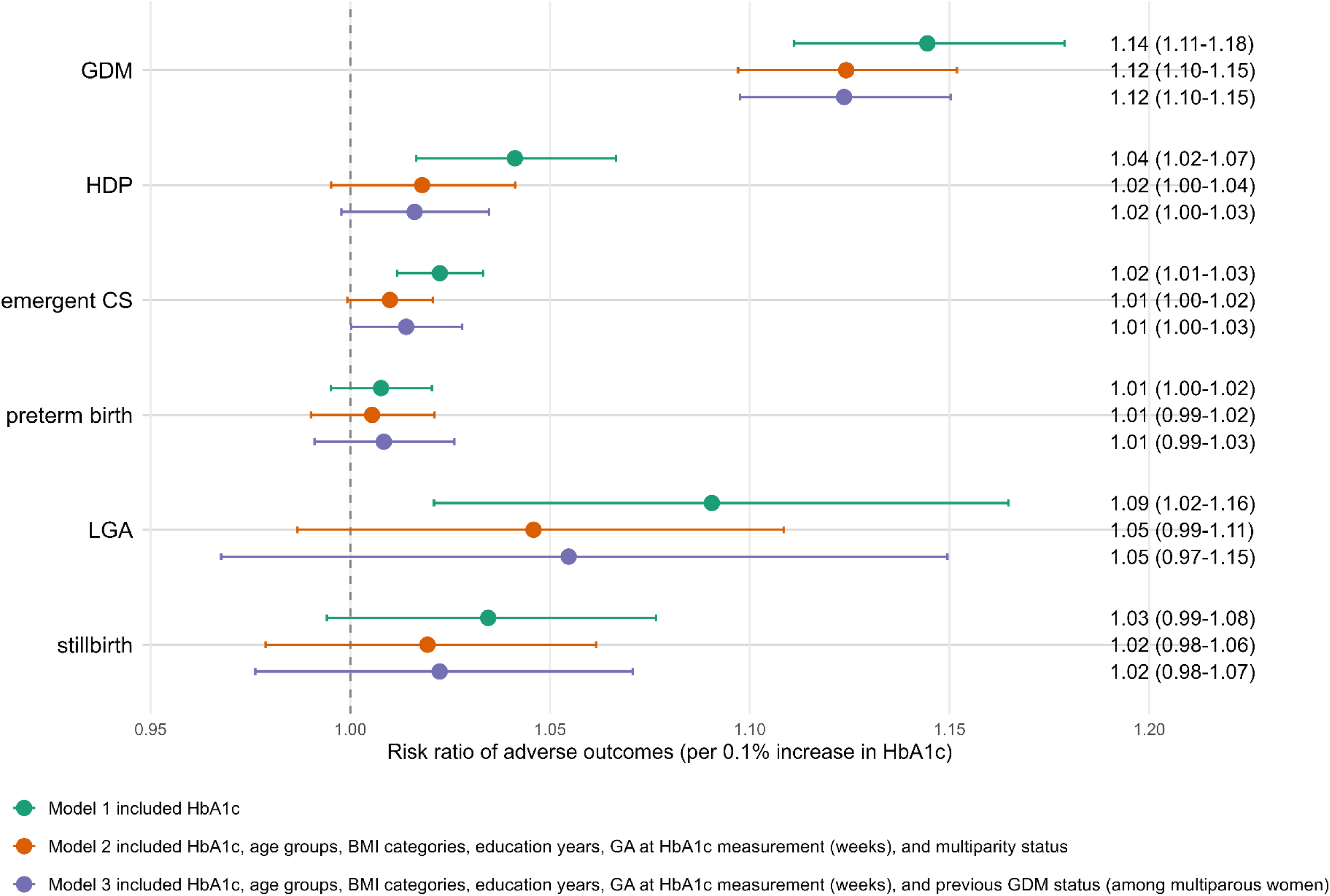
Risk ratios of early HbA1c on GDM and adverse outcomes.

In sensitivity analyses, regressions restricted to those without anemia or to those with normal early HbA1c (< 5.7%) yielded results similar to those of the primary analysis **(Supplemental Table 7** and **Table 8)**; however, among those without anemia, a significant association with emergent Cesarean section was observed in model 2.

Similarly, excluding women whose HbA1c was measured after GA 18 weeks did not change the main findings except for an association with HDP among multiparous participants **(Supplemental Table 9)**. In the analysis using early HbA1c as dichotomous predictors, participants with early HbA1c below the rule-out cutoff (5.0%) had a significantly lower risk of GDM than those above this cutoff whereas those with HbA1c at or above the rule-in cutoff (5.5%) had a significantly elevated risk of GDM **(Supplemental Table 10)**.

The GAM analysis showed nonlinear relationships between early HbA1c and the predicted probabilities of adverse outcomes, with some turning points noted and variations across sites. In the model examining early HbA1c and GDM, slight increases in predicted probabilities were observed around HbA1c 5.4%, the single optimal cutoff we identified, for Zambia, North India, and Pakistan, with a more significant rise around the prediabetes cutoff of 5.7% for Pakistan. In contrast, South India showed a sharp rise in the predicted probability before HbA1c 5.4% **(Supplemental Figure 2)**.

## Discussion

In this study, which used data from a large, multi-country cohort of pregnant women across four LMICs, we found that higher early pregnancy HbA1c was independently associated with an increased risk of GDM after adjusting for maternal age and BMI. Although the overall predictive performance of a single HbA1c cutoff alone was suboptimal, we identified rule-in and rule-out HbA1c cutoffs that could potentially be used to identify pregnant women at higher and lower risk of subsequent GDM.

Consistent with the existing literature, we found that early pregnancy HbA1c is associated with the development of later GDM. In adjusted analyses, each 0.1% increase in early HbA1c is associated with an elevated risk of GDM (RR 1.12, 95% CI 1.10–1.15). These results are consistent with the estimates reported by the STRiDE study, which included 2,100 participants each in Kenya and India and reported adjusted risk ratios of 1.04 (95% CI 1.01–1.07) in India and 1.10 (95% CI 1.08–1.13) in Kenya.[4] While the adjustment strategies differed slightly between studies, both analyses accounted for key maternal characteristics, including age and BMI. Similarly, Hinkle et al. demonstrated a dose-dependent association, with each 0.1% increase in HbA1c measured at GA 8 to 13 weeks associated with a 22% higher risk of subsequent GDM in a US-based cohort.[21] Unlike the STRiDE study and the present analysis, which used HbA1c as a continuous predictor, most prior studies evaluated HbA1c using dichotomous cutoffs, such as 5.7% or 6.0%. A systematic review of 11 studies found increased risks of GDM among participants with an early HbA1c level greater than 5.7%.[26] By examining HbA1c as both a continuous and a dichotomous variable, our study provides a more comprehensive assessment of its association with GDM, thereby adding value to the existing body of literature. Collectively, these findings support an association between early HbA1c and the subsequent development of GDM.

Beyond GDM, associations between early HbA1c and secondary outcomes were largely null in our primary adjusted model. In contrast, previous literature has shown evidence of associations between early HbA1c and other adverse perinatal outcomes, particularly preeclampsia, preterm birth, and LGA [27, 28]; evidence on Cesarean sections is mixed [27, 28], and studies on stillbirth are limited.[27] A meta-analysis of 17 studies found that an early HbA1c above 5.7% was associated with high risks of preeclampsia (RR 2.02, 95% CI 1.53–2.66) and LGA (RR 1.38, 95% CI 1.15–1.66), and the association with preterm birth (RR 1.67, 95%CI 1.39–2.00) was significant regardless of HbA1c cutoffs.[28] However, these findings should be interpreted cautiously. First, most previous research used various prespecified HbA1c cutoffs, which likely affected the effect sizes; second, prior studies on this topic were heterogeneous in their study populations and the timing of HbA1c measurement, etc.[28] Also, there was limited representation of low-resource settings in the existing body of literature. A secondary analysis using the data from the AMANHI cohort in Bangladesh, Pakistan, and Tanzania found that HbA1c ≥6.5% between GA 8 and 20 weeks was associated with higher risks of stillbirth, preterm birth, and Cesarean section, though their higher cutoff limits direct comparability with our analysis.[27] In addition, limited statistical power due to some uncommon secondary outcomes in our sample, such as LGA and stillbirth after 28 weeks, may have partially contributed to our null findings.

Meanwhile, the pathogenesis and mechanisms underlying the associations between early HbA1c and adverse outcomes beyond GDM remain incompletely understood.[28] Evidence suggests a pathway through later GDM, as early HbA1c is associated with GDM, which in turn is linked to other adverse outcomes. On the other hand, GDM-independent mechanisms underlying the association between early HbA1c and adverse outcomes, such as oxidative stress, inflammation, and placental dysfunction, are plausible [28, 29]; further investigation using rigorously designed studies is warranted. In addition, physiological changes in HbA1c during pregnancy complicate the interpretation of studies examining how HbA1c is associated with perinatal outcomes. Pregnancy-related hemodilution and increased red blood cell turnover might lower measured HbA1c levels. The upper limit of the normal range falls from approximately 6.3% before pregnancy to 5.7% in early pregnancy.[30] At the same glycemic levels, HbA1c is lower in pregnancy than in the non-pregnant state. Various hemoglobin variants and anemia also affect HbA1c levels.[31, 32] Iron-deficiency anemia (IDA) elevates HbA1c independent of glycemia: one study found that non-diabetic people with IDA had a mean HbA1c of 5.8% versus 5.3% in healthy adults without IDA, and their HbA1c fell to 5.4% after iron therapy.[33] By contrast, sickle-cell trait and conditions that shorten red-cell lifespans, such as hemolytic anemia, can falsely lower HbA1c levels.[34, 35] In summary, physiologic changes in HbA1c during pregnancy, conditions that affect red blood cell turnover, and the potential development of later GDM complicate the interpretation of the relationship between early HbA1c and adverse perinatal outcomes.

We found that early HbA1c had limited performance as a standalone test for GDM, with sensitivity (46.7%) and specificity (75.8%). Our single optimal cutoff has low sensitivity, consistent with prior studies [14, 36–38], which is plausible, as GDM typically manifests in the second half of pregnancy when rising pregnancy hormones lead to insulin resistance.[39] Early HbA1c levels are unlikely to reflect later pregnancy glycemia of GDM patients.[40] A systematic review by Amaefule et al., which included 23 studies, reached a similar conclusion: no single cutoff for early HbA1c is sufficiently sensitive in predicting GDM, and HbA1c must be paired with a more sensitive test to avoid missing true cases.[14] The core problem is that early HbA1c distributions between women with and without GDM overlap substantially, so any threshold that catches more cases also flags more false positives.[41]

Because early HbA1c has limited performance as a standalone test, researchers have explored a two-threshold approach. In a study in Spain, Benaiges et al. identified a low early HbA1c cutoff to rule out low-risk women and a high cutoff to identify likely GDM, potentially reducing OGTT use without relying on a single imperfect threshold.[38] We applied the two-threshold approach, similar to that used in the STRiDE cohort study, also done in low-resource settings.[4] Our rule-out cutoff of 5.0% achieved a high NPV (96.4%), comparable to STRiDE India’s 4.9% and Kenya’s 5.2%, suggesting that women below this threshold may be candidates for skipping OGTT testing. In our sample, 34.6% fell below the lower rule-out cutoff, compared with 22.0% in the Indian site and 45.3% in the Kenya site from the STRiDE study, indicating a modest potential to reduce OGTT use in our study settings. At the higher end, our cutoff of ≥5.5% corresponded to a very low PPV (12.2%), suggesting substantial false positives among those above the cutoff, which might increase the burden of unnecessary investigation and cause patient anxiety. Thus, while using rule-out and rule-in cutoffs might modestly reduce OGTT testing, women above the higher cutoff and between the two cutoffs would still require an OGTT. It is also worth noting that both the rule-out and rule-in cutoffs we found are below the conventional prediabetes cutoff of 5.7%. Pregnant women who subsequently develop GDM may have normal HbA1c levels in early pregnancy as the underlying insulin resistance has not yet developed, reflecting an inherent biological limitation of using early HbA1c.The overall GDM prevalence in our cohort (6.4%) was lower than the standardized regional estimates reported by the IDF Diabetes Atlas, which may partly reflect differences in population characteristics and possible under-ascertainment.

Key strengths of our study include its large, multi-country sample drawn using standardized protocols and high-quality data from five low-resource settings where evidence on this topic is extremely sparse. However, several important limitations exist. First, we did not adjust for hemoglobin concentration, hemoglobinopathies, or iron status in the main models, even though we excluded participants with severe anemia, which may have affected site-specific findings given the high prevalence of anemia and iron deficiency in our population. We acknowledged the limitation of excluding only women with severe anemia and conducted relevant sensitivity analysis. Second, different methods used to measure HbA1c across sites might introduce bias and cause heterogeneity in findings, while harmonized and standardized procedures and quality assurance programs were in place. Third, excluding data from the Ghana site might have introduced bias and affected the representativeness of our findings; however, data from the two African sites and three Asian sites still provide broad representation of our cohort. our approach to only including pregnancies that have ended after 24 weeks will not allow for assessing associations between early HbA1c and early pregnancy outcomes, such as pregnancy losses before 24 weeks. Fourth, regarding the outcome of stillbirth, we focused on stillbirths that happened after 28 weeks, meaning the stillbirths happening between 24 and 28 weeks were not assessed. We opted to use the World Health Organization definition of 28 weeks as the cutoff for stillbirths.[42]

## Conclusion

Early-pregnancy HbA1c was independently associated with subsequent GDM across five sites in four LMICs but had limited standalone predictive performance. Although early HbA1c is unlikely to serve as a replacement for OGTT, it may have a role in identifying women at particularly low or high risk of GDM. Further studies are needed to determine whether such testing can effectively and efficiently reduce OGTT testing in resource-constrained settings.

## Data Availability

The analytical dataset may be made available upon reasonable request to the corresponding author, Dr. Anne George Cherian, at, and with appropriate IRB approvals from each institution.

## Declarations

Ethics approval and consent to participate: The study protocol has been approved by the relevant institutional review board (IRB) and ethics review committee (ERC) responsible for oversight of the study at each site. All procedures performed in this study involving human participants were in accordance with the ethical standards of the institutional and/or national research committee and with the 1964 Helsinki Declaration and its later amendments or comparable ethical standards. IRB approvals for this research were received from the following ERCs in each country: Ghana (Ghana Health Service ERC (FWA No. 000200025) and the Kintampo Health Research Center Institutional Ethics Committee, FWA No. 00011103), Kenya (KEMRI Scientific and Ethics Review Unit, 04-10-358-4166), Zambia (University of Zambia Biomedical Research Ethics Committee, IRB00001131 of IORG0000774 and UNC Biomedical IRB 356795), India (Office of Research, Christian Medical College, Vellore, India Ethics Committee, IRB14553), India (Ethics Review Committee, Society for Applied Studies, New Delhi, SAS/ERC/ReMAPP Study/2022; Department of Health Research, EC/NEW/INST/2022/DL/0140), Pakistan (National Institutes of Health - Health Research Institute, National Bioethics Committee Ref: No.4-87/NBC-962/23/593 and Aga Khan University Ethics Review Committee, 2022-7197-21350), and the United States (Columbia University IRB IRB-AAAU7504; The George Washington University IRB NCR224396; Harvard University IRB IRB-23-1093).

Informed consent to participate was obtained from all the participants in the study.

## Consent for publication

Not applicable

## Availability of data and materials

The analytical dataset may be made available by reasonable request to the corresponding author, Dr. Anne George Cherian, at, and with appropriate IRB approvals from each institution.

## Authors’ contributions

Conceptualization: MIN, QP, ERS, AGC. Study design: BV, MIN, SJB, NS, SM, VA, FAA, ERS, CM, AGC. Funding acquisition: MIN, SJB, SM, ZH, VA, FAA, ERS, CM, AGC. Data acquisition: BV, BLF, MIN, SJB, JAG, NS, SM, ZH, VA, FAA, ERS, CM, MBS, MPK, AGC. Data management: EMO, MBS, MPK. Data analysis: WCY, QP. Writing - original draft: WCY, SSJ. Writing - review and editing: all authors. All authors have read and approved the final manuscript.

## Acknowledgements

We are grateful to all the women and their families who participated in this study across Kenya, India, Pakistan, and Zambia. Their willingness to contribute their time and data made this research possible. We thank the field teams, research nurses, midwives, laboratory staff, and data managers at each of the five study sites - Christian Medical College (Vellore, South India), Society for Applied Studies (North India), Aga Khan University (Karachi, Pakistan), Kenya Medical Research Institute (Kisumu, Kenya), and UNC Global Projects Zambia (Lusaka, Zambia) for their dedication to data collection and participant care throughout the study. We also thank the broader PRISMA-MNH study team members who contributed to study implementation but are not listed as authors on this manuscript.

This work was supported by the Gates Foundation grant numbers [INV-047400 to KPA, CTA, and SN; INV-057219 to VA; INV-043092 to SB; INV-057220 to ZH; INV-016221 to MPK; INV-057222 to WM; INV-041999 to ERS; INV-060797 to CNM; and INV-057223 to SM]. The conclusions and opinions expressed in this work are those of the author(s) alone and shall not be attributed to the Foundation. Under the grant conditions of the Foundation, a Creative Commons Attribution 4.0 License has already been assigned to the Author Accepted Manuscript version that might arise from this submission. Please note works submitted as a preprint have not undergone a peer review process.

## PRISMA Consortium Collaborator Statement

The Pregnancy Risk, Infant Surveillance, and Measurement Alliance (PRISMA) Consortium includes the following institutions and members: **Aga Khan University** (Zahra Hoodbhoy, Fyezah Jehan, Amna Khan, Muhammad Imran Nisar, Asad Sheikh, Shayan Khakwani, Kinza Farooqui, Danish Hudani, Aamir Abbas, Muhammad Kashif, Nida Yazdani); **Beth Israel Deaconess Medical Center** (Blair J. Wylie); **Christian Medical College, Vellore** (Anne George Cherian, Santosh Joseph Benjamin, James A, Indhumathi, Daniel Jebakumar, Lydia Vasanth, Venkata Raghava Mohan, Vijayalekshmi B, Jayakumar Amirtharaj G, Pamela Christudas, John Jude Antony Prakash, Dhanalakshmi Solaimali, Rani Diana Sahni, John Fletcher, Asha Mary Abraham,Priya Abraham, Rajesh Kannangai, Divya M, Manish Kumar, Mintoo M Tergestina, Sebin George Abraham, Beena Koshy, Molly Jacob); **Dr. Dang’s Laboratory** (Leena Chatterjee, Arjun Dang, Manavi Dang, R Venketeshwar); **Gates Foundation** (Laura M. Lamberti); **The George Washington University Milken Institute School of Public Health** (Nazia Binte Ali, Sasha G. Baumann, Emma Cook, Bethany Freeman, Xinyi Li, Casey Kalman, Jaime Marquis, Jamie Minchin, Christopher N. Mores, Erin M. Oakley, Savannah O’Malley, Qing Pan, Abigale Proctor, Jennifer Seager, Alyssa Shapiro, Emily R. Smith, Ziwei Song, Precious Williams, Wen-Chien Yang); **Harvard T.H. Chan School of Public Health** (Christopher R. Sudfeld); **Kenya Medical Research Institute and Liverpool School of Tropical Medicine** (Victor Akelo, Florence Aweyo, Kephas Otieno, Harun Owuor, Dickens Onyango, Caleb Sagam, Feiko ter Kuile, Joyce Were, Zacchaeus Were, Dickson Gethi, Dorothy Lynda Achieng, Kevin Kasadhe, Edwin Kiplelgo); **Kintampo Health Research Centre** (Irene Apewe Adjei, Ken Ae-Ngibise, Veronica Agyemang, Japhet Anim, Kwaku Poku Asante, Ellen Boamah-Kaali, Richard Boakye, Stephaney Gyasse, Sam Newton, Eliezer Odei-Lartey, Samuel Addo Oppong, Richard Tetteh); **Society for Applied Studies** (Sarmila Mazumder, Neeraj Sharma, Arun Singh Jadaun, Rupa Talukdar, Mrinal Kishore, Dinesh Kumar Dhingra, Meghna Singh, Munita Jat, Kamal Kant, Soumya R. Nayak); University of Alabama at Birmingham School of Medicine (Lynda Ugwu); **University of North Carolina—Global Projects Zambia** (Margaret P. Kasaro, Felistas Mbewe, Twaambo Munaumba, Humphrey Mwape, Augustine Tunga); **University of North Carolina at Chapel Hill** (M. Bridget Spelke, Jeffrey S.A. Stringer); **University of Zambia** (Wilbroad Mutale, Mutale Sampa, Bellington Vwalika); and **VITAL Pakistan Trust** (Kinza Farooqui, Farzana Shaheen).

## References

1. Kjos SL, Buchanan TA. Gestational diabetes mellitus. N Engl J Med. 1999;341:1749– 56.

2. Saeedi M, Cao Y, Fadl H, Gustafson H, Simmons D. Increasing prevalence of gestational diabetes mellitus when implementing the IADPSG criteria: A systematic review and meta-analysis. Diabetes Res Clin Pract. 2021;172:108642.

3. Wang H, Li N, Chivese T, Werfalli M, Sun H, Yuen L, et al. Diabetes Atlas: estimation of global and regional gestational diabetes mellitus prevalence for 2021 by International Association of Diabetes in Pregnancy Study Group’s criteria. Diabetes Res Clin Pract. 2022;183:109050.

4. Saravanan P, Deepa M, Ahmed Z, Ram U, Surapaneni T, Kallur SD, et al. Early pregnancy HbA1c as the first screening test for gestational diabetes: results from three prospective cohorts. Lancet Diabetes Endocrinol. 2024;12:535–44.

5. Ye W, Luo C, Huang J, Li C, Liu Z, Liu F. Gestational diabetes mellitus and adverse pregnancy outcomes: systematic review and meta-analysis. BMJ. 2022;377:e067946.

6. Lowe LP, Metzger BE, Dyer AR, Lowe J, McCance DR, Lappin TRJ, et al. Hyperglycemia and Adverse Pregnancy Outcome (HAPO) Study: associations of maternal A1C and glucose with pregnancy outcomes. Diabetes Care. 2012;35:574–80.

7. Wang H, Li N, Chivese T, Werfalli M, Sun H, Yuen L, et al. IDF Diabetes Atlas: estimation of global and regional gestational diabetes mellitus prevalence for 2021 by International Association of Diabetes in Pregnancy Study Group’s criteria. Diabetes Res Clin Pract. 2022;183:109050.

8. Diagnostic criteria and classification of hyperglycaemia first detected in pregnancy: a World Health Organization Guideline. Diabetes Res Clin Pract. 2014;103:341–63.

9. Putoto G, Somigliana E, Olivo F, Ponte S, Koroma MM, Citterio F, et al. A simplified diagnostic work-up for the detection of gestational diabetes mellitus in low resources settings: achievements and challenges. Arch Gynecol Obstet. 2020;302:1127–34.

10. Utz B, De Brouwere V. “Why screen if we cannot follow-up and manage?” Challenges for gestational diabetes screening and management in low and lower-middle income countries: results of a cross-sectional survey. BMC Pregnancy Childbirth. 2016;16:341.

11. Hinneh T, Jahn A, Agbozo F. Barriers to screening, diagnosis and management of hyperglycaemia in pregnancy in Africa: a systematic review. Int Health. 2022;14:211– 21.

12. Bhavadharini B, Uma R, Saravanan P, Mohan V. Screening and diagnosis of gestational diabetes mellitus - relevance to low and middle income countries. Clin Diabetes Endocrinol. 2016;2:13.

13. World Health Organization. WHO recommendations on antenatal care for a positive pregnancy experience. Geneva, Switzerland: World Health Organization; 2016.

14. Amaefule CE, Sasitharan A, Kalra P, Iliodromoti S, Huda MSB, Rogozinska E, et al. The accuracy of haemoglobin A1c as a screening and diagnostic test for gestational diabetes: a systematic review and meta-analysis of test accuracy studies. Curr Opin Obstet Gynecol. 2020;32:322–34.

15. Renz PB, Chume FC, Timm JRT, Pimentel AL, Camargo JL. Diagnostic accuracy of glycated hemoglobin for gestational diabetes mellitus: a systematic review and meta-analysis. Clin Chem Lab Med. 2019;57:1435–49.

16. Herman WH, Cohen RM. Racial and Ethnic Differences in the Relationship between HbA1c and Blood Glucose: Implications for the Diagnosis of Diabetes. J Clin Endocrinol Metab. 2012;97:1067–72.

17. Cavagnolli G, Pimentel AL, Freitas PAC, Gross JL, Camargo JL. Effect of ethnicity on HbA1c levels in individuals without diabetes: Systematic review and meta-analysis. PLoS One. 2017;12:e0171315.

18. Pregnancy Risk, Infant Surveillance, and Measurement Alliance (PRISMA) Investigators. Pregnancy Risk, Infant Surveillance, and Measurement Alliance (PRISMA) Maternal and Newborn Health Study: protocol for a multisite, prospective, open cohort study of pregnancy and postpartum health outcomes in South Asia and sub-Saharan Africa. BMJ Open. 2026;16:e104512.

19. Smith ER, Hoodbhoy Z, Hotwani A, Jehan F, Khan A, Nisar I, et al. Protocol for the Redefining Maternal Anemia in Pregnancy and Postpartum (ReMAPP) study: A multisite, international, population-based cohort study to establish global hemoglobin thresholds for maternal anemia. PLoS One. 2025;20:e0321943.

20. Villar J, Cheikh Ismail L, Victora CG, Ohuma EO, Bertino E, Altman DG, et al. International standards for newborn weight, length, and head circumference by gestational age and sex: the Newborn Cross-Sectional Study of the INTERGROWTH-21st Project. Lancet. 2014;384:857–68.

21. Hinkle SN, Tsai MY, Rawal S, Albert PS, Zhang C. HbA1c measured in the first trimester of pregnancy and the association with gestational diabetes. Sci Rep. 2018;8:12249.

22. Lamain-de Ruiter M, Kwee A, Naaktgeboren CA, Franx A, Moons KGM, Koster MPH. Prediction models for the risk of gestational diabetes: a systematic review. Diagn Progn Res. 2017;1:3.

23. Zhang Y, Xiao C-M, Zhang Y, Chen Q, Zhang X-Q, Li X-F, et al. Factors Associated with Gestational Diabetes Mellitus: A Meta-Analysis. Journal of Diabetes Research. 2021;2021:6692695.

24. Wang J-W, Wang Q, Wang X-Q, Wang M, Cao S-S, Wang J-N. Association between maternal education level and gestational diabetes mellitus: a meta-analysis. J Matern Fetal Neonatal Med. 2021;34:580–7.

25. Schwarzer G. meta: An R Package for Meta-Analysis. R News. 2007;7:40–5.

26. Kattini R, Hummelen R, Kelly L. Early gestational diabetes mellitus screening with glycated hemoglobin: A systematic review. J Obstet Gynaecol Can. 2020;42:1379–84.

27. Nisar MI, Das S, Khanam R, Khalid J, Chetia S, Hasan T, et al. Early to mid-pregnancy HbA1c levels and its association with adverse pregnancy outcomes in three low middle-income countries in Asia and Sub-Saharan Africa. BMC Pregnancy Childbirth. 2024;24:66.

28. Mañé L, Navarro H, Pedro-Botet J, Chillarón JJ, Ballesta S, Payà A, et al. Early HbA1c Levels as a Predictor of Adverse Obstetric Outcomes: A Systematic Review and Meta-Analysis. J Clin Med. 2024;13:1732.

29. Cohen AL, Wenger JB, James-Todd T, Lamparello BM, Halprin E, Serdy S, et al. The association of circulating angiogenic factors and HbA1c with the risk of preeclampsia in women with preexisting diabetes. Hypertension in Pregnancy: 2014;33:81–92.

30. Nielsen LR, Ekbom P, Damm P, Glümer C, Frandsen MM, Jensen DM, et al. HbA1c levels are significantly lower in early and late pregnancy. Diabetes Care. 2004;27:1200– 1.

31. English E, Idris I, Smith G, Dhatariya K, Kilpatrick ES, John WG. The effect of anaemia and abnormalities of erythrocyte indices on HbA1c analysis: a systematic review. Diabetologia. 2015;58:1409–21.

32. Little RR, Roberts WL. A review of variant hemoglobins interfering with hemoglobin A1c measurement. J Diabetes Sci Technol. 2009;3:446–51.

33. Alzahrani BA, Salamatullah HK, Alsharm FS, Baljoon JM, Abukhodair AO, Ahmed ME, et al. The effect of different types of anemia on HbA1c levels in non-diabetics. BMC Endocr Disord. 2023;23:24.

34. Lacy ME, Wellenius GA, Sumner AE, Correa A, Carnethon MR, Liem RI, et al. Association of sickle cell trait with hemoglobin A1c in African Americans. JAMA. 2017;317:507–15.

35. Jiao Y, Okumiya T, Saibara T, Park K, Sasaki M. Abnormally decreased HbA1c can be assessed with erythrocyte creatine in patients with a shortened erythrocyte age. Diabetes Care. 1998;21:1732–5.

36. Immanuel J, Simmons D, Desoye G, Corcoy R, Adelantado JM, Devlieger R, et al. Performance of early pregnancy HbA1c for predicting gestational diabetes mellitus and adverse pregnancy outcomes in obese European women. Diabetes Res Clin Pract. 2020;168:108378.

37. Osmundson SS, Zhao BS, Kunz L, Wang E, Popat R, Nimbal VC, et al. First trimester hemoglobin A1c prediction of gestational diabetes. Am J Perinatol. 2016;33:977–82.

38. Benaiges D, Flores-Le Roux JA, Marcelo I, Mañé L, Rodríguez M, Navarro X, et al. Is first-trimester HbA1c useful in the diagnosis of gestational diabetes? Diabetes Res Clin Pract. 2017;133:85–91.

39. Catalano PM. Trying to understand gestational diabetes. Diabet Med. 2014;31:273– 81.

40. Edelson PK, James KE, Leong A, Arenas J, Cayford M, Callahan MJ, et al. Longitudinal Changes in the Relationship Between Hemoglobin A1c and Glucose Tolerance Across Pregnancy and Postpartum. J Clin Endocrinol Metab. 2020;105:e1999–2007.

41. Agarwal MM, Dhatt GS, Punnose J, Koster G. Gestational diabetes: a reappraisal of HbA1c as a screening test. Acta Obstet Gynecol Scand. 2005;84:1159–63.

42. World Health Organization. Making every baby count: audit and review of stillbirths and neonatal deaths. World Health Organization. World Health Organization; 2016.

